# *ZFHX3* variants cause childhood partial epilepsy and infantile spasms with favorable outcomes

**DOI:** 10.1101/2023.07.16.23292551

**Authors:** Ming-Feng He, Li-Hong Liu, Sheng Luo, Juan Wang, Jia-Jun Guo, Peng-Yu Wang, Qiong-Xiang Zhai, Su-Li He, Dong-Fang Zou, Xiao-Rong Liu, Bing-Mei Li, Hai-Yan Ma, Jing-Da Qiao, Peng Zhou, Na He, Yong-Hong Yi, Wei-Ping Liao

## Abstract

**Background:** The *ZFHX3* gene plays vital roles in embryonic development, cell proliferation, neuronal differentiation, and neuronal death. This study aims to explore the relationship between *ZFHX3* variants and epilepsy.

**Methods:** Whole-exome sequencing was performed in a cohort of 378 patients with partial (focal) epilepsy. A *Drosophila Zfh2* knockdown model was used to validate the association between *ZFHX3* and epilepsy.

**Results:** Compound heterozygous *ZFHX3* variants were identified in eight unrelated cases. The burden of *ZFHX3* variants was significantly higher in the case cohort, shown by multiple/specific statistical analyses. In *Zfh2* knockdown flies, the incidence and duration of seizure-like behavior were significantly greater than those in the controls. The *Zfh2* knockdown flies exhibited more firing in excitatory neurons. All patients presented partial seizures. The five patients with variants in the C-terminus/N-terminus presented mild partial epilepsy. The other three patients included one who experienced frequent nonconvulsive status epilepticus and two who had early spasms. These three patients had also neurodevelopmental abnormalities and were diagnosed as developmental epileptic encephalopathy (DEE), but achieved seizure-free after antiepileptic-drug treatment without adrenocorticotropic-hormone/steroids. The analyses of temporal expression (genetic dependent stages) indicated that *ZFHX3* orthologs were highly expressed in the embryonic stage and decreased dramatically after birth.

**Conclusion:** *ZFHX3* is a novel causative gene of childhood partial epilepsy and DEE. The patients of infantile spasms achieved seizure-free after treatment without adrenocorticotropic-hormone/steroids implies a significance of genetic diagnosis in precise treatment. The genetic dependent stage provided an insight into the underlying mechanism of the evolutional course of illness.

**WHAT IS ALREADY KNOWN ON THIS TOPIC:** The ZFHX3 protein plays an essential role in neurodevelopment. The relationship between *ZFHX3* variants and human diseases remains unknown.

**WHAT THIS STUDY ADDS:** Eight pairs of compound heterozygous *ZFHX3* variants were identified in eight unrelated patients with partial epilepsy, including two who evolved from early spasms.

**HOW THIS STUDY MIGHT AFFECT RESEARCH, PRACTICE OR POLICY:** The *ZFHX3* gene is a novel pathogenic gene of childhood partial epilepsy and developmental epileptic encephalopathy. The development-dependent expression pattern of *ZFHX3* explains the evolutional course of the illness, potentially being helpful in the management of the patients.

## INTRODUCTION

Epilepsy is a common neurological disorder with age-dependent seizures. In children, the prevalence of epilepsy ranges from 0.5% to 1%,[1] and approximately 68% of childhood patients experience focal seizures.[2] In infants, epilepsy affects approximately 0.7% of the population,[3] and 10% of infantile patients with epilepsy are diagnosed as infantile spasms that is a common form of developmental epileptic encephalopathy (DEE).[4] Genetic factors are believed to be the main cause of epilepsy, accounting for approximately eighty percent of cases.[5] Previously, a number of causative genes were identified in children with partial (focal) epilepsy, such as *DEPDC5*,[6] *GRIN2A*,[7] *UNC13B*,[8] *HCFC1*,[9] *LAMA5*,[10] *BCOR*,[11] *CELSR1*,[12] *BRWD3*,[13] *FAT1*,[14] and *ELP4*.[15] The genes associated with infantile spasms include *KCNQ2*,[16] *KCNT1*,[17] *GRIN2B*,[18] *STXBP1*,[19] *TBC1D24*,[20] *SCN1A*,[21] *CDKL5*,[22] and *ARX*.[23] However, the etiology in most patients with childhood partial epilepsy and/or infantile spasms remains to be elucidated.

In this study, we conducted trio-based whole-exome sequencing (WES) in a cohort of 378 unrelated children with partial epilepsy. Compound heterozygous variants of *ZFHX3* were identified in eight unrelated patients with partial epilepsy, including one with frequent nonconvulsive status epilepticus and two who evolved from early spasms; the three patients with severe seizures also had neurodevelopmental abnormalities and were diagnosed as DEE early. However, all patients became seizure-free. A *Drosophila* model with *Zfh2* knockdown was established to investigate the association between *ZFHX3* and epilepsy. To explore the underlying mechanism of favorable outcomes, the genetic-dependent stages (GDS)[14, 24] of *ZFHX3* orthologs were investigated, for which the gene expression in flies and mice was determined by RT-qPCR and that in humans was analyzed by using data from the Brainspan database. This study suggested that the *ZFHX3* gene is a novel causative gene of partial epilepsy of childhood and DEE. The development-dependent expression pattern of *ZFHX3* explains the evolutional course of the illness.

## MATERIALS AND METHODS

### Patients

Patients were identified in a cohort of 378 unrelated children with partial epilepsy without acquired causes who were recruited from five hospitals from 2019 to 2022, including the Second Affiliated Hospital of Guangzhou Medical University, Guangdong General Hospital, Shenzhen Children’s Hospital, Shantou Chaonan Minsheng Hospital, and the Affiliated Brain Hospital of Nanjing Medical University. The clinical information of the affected individuals was collected, including sex, seizure onset age, seizure types and frequency, response to antiepileptic drugs (AEDs), family history, and general and neurological examinations. Long-term video-electroencephalography (EEG) was performed to monitor epileptic discharges. Brain magnetic resonance imaging (MRI) scans were performed to detect structural abnormalities. Epileptic seizures and epilepsy syndromes were diagnosed according to the criteria of the Commission on Classification and Terminology of the ILAE (1989, 2001, 2010, 2017, and 2022).

### Trio-based whole-exome sequencing and genetic analysis

Genomic DNA was extracted from blood samples of the probands and their parents (trios) and other available family members by using the Flexi Gene DNA Kit (Qiagen, Hilden, Germany). Trio-based WES was performed with NextSeq500 sequencing instruments (Illumina, San Diego, CA, USA). Detailed sequencing methods were described in our previous studies.[8] A case-by-case analytical approach was adopted to identify candidate causative variants in each trio.[8] Primarily, the rare variants were prioritized with a minor allele frequency (MAF) < 0.005 in the gnomAD database gnomad.broadinstitute.org). Then, potentially pathogenic variants were retained, including frameshift, nonsense, canonical splice site, initiation codon, in-frame variants, missense, and synonymous variants predicted to impact splicing. Last and importantly, possibly disease-causing variants in each individual were screened under five models: 1) epilepsy-associated gene; 2) de novo variant dominant; 3) autosomal recessive inheritance, including homozygous and compound heterozygous variants; 4) X-linked; and 5) cosegregation analysis, if available. To identify novel potential epilepsy genes, genes with recurrently identified de novo, biallelic, hemizygous, and cosegregated variants were selected for further studies. *ZFHX3* appeared to be a candidate gene associated with recurrent compound heterozygous variants in this cohort. All the *ZFHX3* variants identified in this study were validated by Sanger sequencing and annotated based on the transcript NM_006885.4.

### Analysis of the burden of variants

Three specific statistical methods were used to analyze the association between *ZFHX3* and epilepsy incidence, namely, recessive burden analysis,[25] aggregate frequency of variants,[26] and frequency of compound heterozygous variants.

For the burden of recessive variants, the *P* value was calculated as [1-cumulative binomial probability [(n-1, N, R)] according to a previous report,[25] where n is the observed biallelic variant number for *ZFHX3*, N is the number of trios (378 in this cohort), and R is the rate of *ZFHX3* variants by chance in populations. Considering that all patients were Han Chinese, the cutoff was set according to the MAF in the ExAC-East Asian population. The Bonferroni correction was used to adjust the *P* value.

For aggregate frequency analysis, the frequencies of identified variants between the case cohort and the controls were compared, including general and East Asian populations in the gnomAD database, general populations in the ExAC database, and the 33,444 persons without known neuropsychiatric conditions in the Epi25 WES Browser (https://epi25.broadinstitute.org/).

For the control of compound heterozygous variants, we established a cohort of 1942 asymptomatic parents from trios, in whom the compound heterozygous variants were identified by detecting one of the paired variants in the child, based on the fact that one of the paired variants in a parent would transmit to the child. The frequency of identified compound heterozygous *ZFHX3* variants in the case cohort was compared with that in the controls, including the 1942 asymptomatic parent controls and the variant co-occurrence data from gnomAD.[27]

### Molecular structural analysis

Protein modeling was performed to predict the effects of missense variants on the molecular structure by using the I-TASSER tool (https://zhanggroup.org/I-TASSER/). Protein structures were visualized and analyzed by using the PyMOL Molecular Graphics System (version 2.5; Schrödinger, LLC; New York, USA). Changes in the protein stability of the missense variants were predicted by using the I-Mutant Suite (https://folding.biofold.org/cgi-bin/i-mutant2.0.cgi).

### *Drosophila* experiments

Flies were reared at 25°C and 60-70% humidity with a standard cornmeal diet under a 12:12 h light and dark cycle. The *UAS-Zfh2-RNAi* line (CG1449, TH01656.N) was purchased from the TsingHua Fly Center. The *tub-Gal4* driver line and *Canton-s* line were donated by Prof. Liu Ji-Yong (Guangzhou Medical University, Guangzhou, China). *Zfh2* knockdown flies were generated by crossing the *UAS-Zfh2-RNAi* line with the *tub-Gal4* driver line. *Canton-s* was the wild-type line. The knockdown efficiency was detected by using reverse transcription quantitative PCR (RT-qPCR).

To evaluate the role of Z*fh2* deficiency in development, the body length of the fly larvae was assessed as described in our previous study.[28] A bang sensitivity assay was conducted on flies 3-5 days after eclosion to evaluate the seizure-like behavior.[8] The duration and percentage of seizure-like behavior were recorded as described in our previous study.[8] To determine the impact of *Zfh2* knockdown on neuronal excitability, the electrophysiological activity of projection neurons was recorded.[29] Spontaneous activity was assessed using a 700B amplifier, 1440B Digital Analog converter, and pClamp 10.5 software (Molecular Devices, San Jose, CA, USA). A cell with an access resistance of <30 MΩ was used for analysis. Spontaneous EPSP (sEPSP) data > 1 mV were analyzed by Mini Analysis software.[8]

### Assessment of the *ZFHX3* ortholog expression profile

The mRNA expression levels of *ZFHX3* orthologs in different developmental stages in flies and mice were determined by RT-qPCR. For flies, the whole mRNA was extracted in five developmental stages, including third instar larvae, pupae, early adult (day 1), middle adult (day 5), and later adult (day 10). For mice, the mRNA of the frontal cortex was extracted in eight developmental stages, including the fetus, neonate (1 day), infant (1 week), toddle-period (2 weeks), preschool (4 weeks), juvenile-adult (10 weeks), middle-age (15 weeks), and old-age (32 weeks). The sequences of the primers used in this study are listed in **Table S1**. Total RNA was extracted by using the HiPure Universal RNA Mini Kit (Magen Biotechnology, Guangzhou, China). Reverse transcription was performed with the HiScript III RT SuperMix for qPCR (+gDNA wiper) (Vazyme, Nanjing, China) kit, and qPCR was subsequently performed with Taq Pro Universal SYBR qPCR Master Mix (Vazyme, Nanjing, China) and the LightCycler 480 System (Roche) tool.

Human RNA-seq data at different developmental stages (from 8 postconceptional weeks to 40 years) for multiple brain areas were obtained from the Brainspan database (http://www.brainspan.org/). RNA expression was normalized to the reads per kilobase million (RPKM) value. The temporal expression curve was fitted by third-order polynomial least squares to interpret the expression pattern of *ZFHX3* by GraphPad Prism 9.

### Statistical analysis

R statistical software (v4.0.2) and GraphPad Prism 9 were used for statistical analysis. All the quantitative data are presented as the mean ± standard error of the mean (SEM). The number and aggregated frequencies of *ZFHX3* variants in this cohort and controls were compared by a two-tailed Fisher’s exact test. Student’s t test was used to compare two independent samples, and the Mann-Whitney test was used to assess nonparametric data. The *P* value < 0.05 was considered statistically significant.

## RESULTS

### Identification of *ZFHX3* variants

Compound heterozygous variants in *ZFHX3* were identified in eight unrelated cases with partial epilepsy (**Table 1 and Figure 1**). The eight pairs of compound heterozygous variants consisted of eleven missense variants, one frameshift truncation, and one in-frame deletion. The missense variant p.Ser3482Ile was recurrently identified in two cases (Cases 7 & 8), and the variant p.Pro3618Gln was recurrently identified in three cases (Cases 2, 5, & 8). All of the *ZFHX3* compound heterozygous variants were inherited from their asymptomatic parents, consistent with the Mendelian autosomal recessive inheritance pattern.

**Figure 1.**
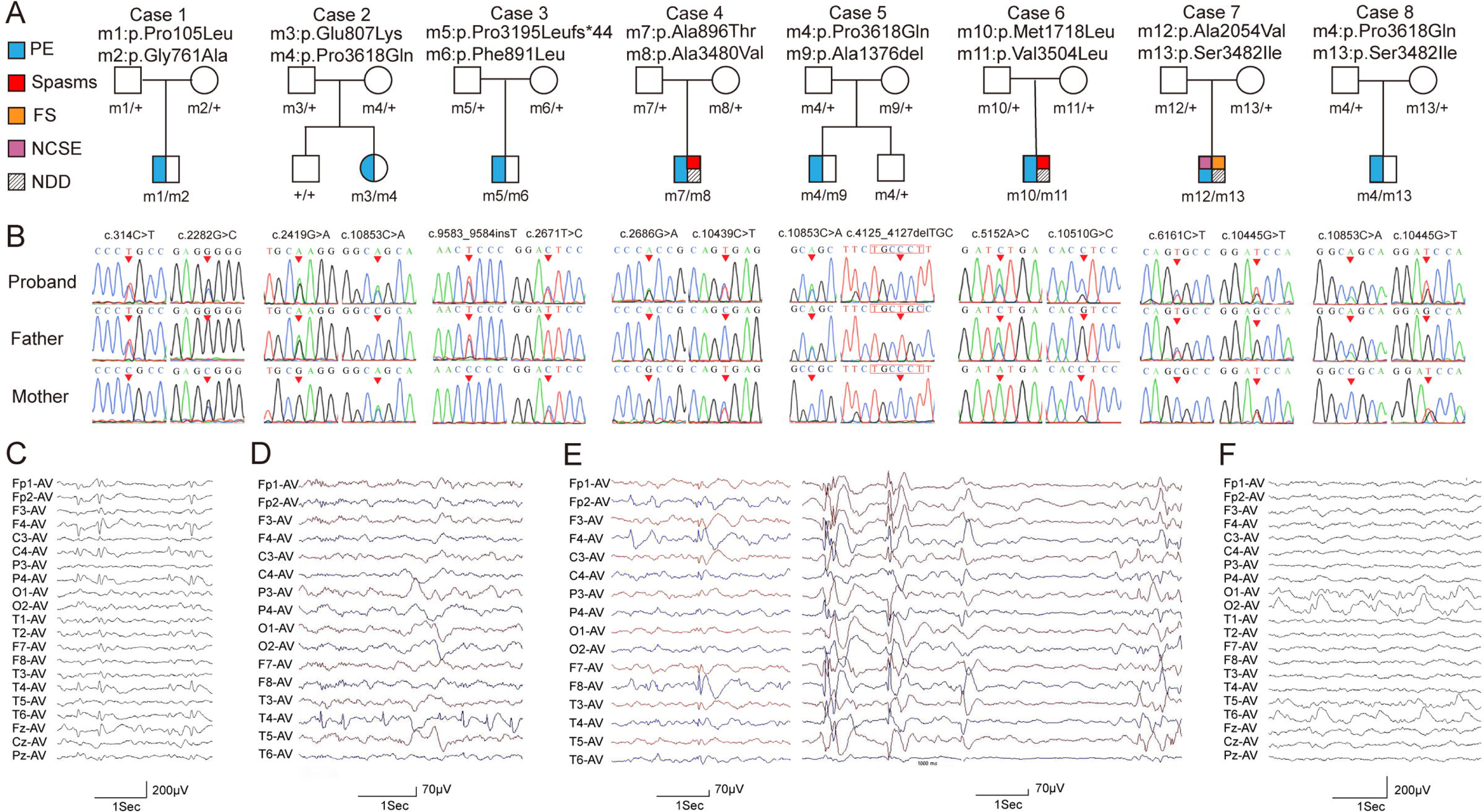
Genetic data and representative EEG recordings of the cases with *ZFHX3* variants. (A) Pedigrees of eight cases with compound heterozygous *ZFHX3* variants and their corresponding phenotypes. PE, partial epilepsy; FS, febrile seizures; NCSE, nonconvulsive status epilepticus; NDD, neurodevelopmental delay. (B) DNA sequencing chromatogram of *ZFHX3* variants. Arrows indicate the position of the variants. (C) EEG of case 2 showed right frontal-centro-temporal sharp-slow waves. (D) EEG of case 5 showed right temporal spike-slow waves. (E) EEG of case 6 showed right frontal spike-slow waves (left) and generalized poly-spike-slow waves (right). (F) EEG of case 8 showed bilateral occipital-temporal sharp-slow waves.

**Table 1.** Clinical features of the individuals with *ZFHX3* variants.

| Case | Variants<br>(NM_006885.4) | Sex/Onset age | Sz course | Sz-free<br>duration | Effective<br>AEDs | EEG | Development | Diagnosis |
| --- | --- | --- | --- | --- | --- | --- | --- | --- |
| Case 1 | c.314C>T/p.Pro105Leu<br>c.2282G>C/p.Gly761Ala | M/childhood | sGTCS, 1-2 times/yr | 1 yr | LEV | Bilateral spike-slow waves with frontal dominance | Normal | PE |
| Case 2 | c.2419G>A/p.Glu807Lys<br>c.10853C>A/p.Pro3618Gln | F/childhood | sGTCS, 1-5 times/mo | 1.5 yr | LTG | Right frontal-centro-temporal sharp-slow waves | Normal | PE |
| Case 3 | c.2671T>C/p.Phe891Leu<br>c.9583_9584insT/p.Pro3195LeufsTer44 | M/toddler | CPS, 2-3 times/yr | 1 yr | OXC | Spike-slow waves in bilateral frontal-central area with right dominance | Normal | PE |
| Case 4 | c.2686G>A/p.Ala896Thr<br>c.10439C>T/p.Ala3480Val | M/infant | Spasms, CPS >10 times/day | 2 yr | LEV, LTG | Poly-spike-slow waves | GDD | DEE |
| Case 5 | c.4125_4127del/p.Ala1376del<br>c.10853C>A/p.Pro3618Gln | M/childhood | sGTCS, 3 times for 2 yrs;<br>CPS, twice at for 1 yr | 5 yr | OXC, VPA | Right temporal spike-slow waves | Normal | PE |
| Case 6 | c.5152A>C/p.Met1718Leu<br>c.10510G>C/p.Val3504Leu | M/toddler | Spasms 5-7 times/day for 4 mos; then CPS 0-10 times/day | 9 yr | VPA, LEV, LTG | Right frontal spike-slow waves with tendency of generalization | GDD | DEE |
| Case 7 | c.6161C>T/p.Ala2054Val<br>c.10445G>T/p.Ser3482Ile | M/infant | FS, 3 times; sGTCS and CPS twice/week; NCSE 2-3 times for 1 yr | 6 mo | VPA, PER, LEV | Left frontal, temporal, and occipital 1.5-2 Hz spike-slow waves; Diffused 1.5-2.5 Hz spike-slow waves | GDD | DEE |
| Case 8 | c.10445G>T/p.Ser3482Ile<br>c.10853C>A/p.Pro3618Gln | F/childhood | sGTCS, 2 times for 2 yr | 1.5 yr | TPM | Bilateral occipital-temporal sharp-slow waves | Normal | PE |

Abbreviations: AEDs, antiepileptic drugs; CPS, complex partial seizures; DEE, developmental epileptic encephalopathy; GDD, global developmental delay; EEG, electroencephalogram; FS, febrile seizures; LEV, levetiracetam; LTG, lamotrigine; NCSE, nonconvulsive status epilepticus; OXC, oxcarbazepine; PE, partial epilepsy; PER, perampanel; Sz, seizures; sGTCS, secondary generalized tonic-clonic seizures; TPM, topamax; VPA, valproate; mo, month; yr, year

The identified variants were absent or presented with low frequencies in the gnomAD database (MAF < 0.005) (**Table S2**). Six variants were not present in the normal control of the Epi25 WES Browser, while the other seven variants presented with extremely low frequencies (MAF < 0.0005). None of the variants were presented as homozygous states in gnomAD.

When the burden of recessive variants was analyzed,[25] the number of recessive *ZFHX3* variants identified in this cohort was significantly greater than the expected number by chance in the East Asian population (ExAC-MAF < 0.001; *P* = 8.52 × 10^−7^, *P* = 0.017 after Bonferroni correction). The aggregate frequency of variants identified in the case cohort was significantly higher than that in the controls, including the controls of the gnomAD-all population (*P* = 2.30×10^-13^), the controls of the gnomAD-East Asian population (*P* = 4.36×10^−2^), the general population of the ExAC database (*P* = 2.34×10^-13^), and the normal controls of the Epi25 WES Browser (*P* = 3.56×10^-18^) (**Table S2**). The eight pairs of compound heterozygous *ZFHX3* variants did not co-occur in gnomAD, and the frequency of compound heterozygous *ZFHX3* variants in the case cohort was significantly higher than that in the controls, including the control cohort of 1942 asymptomatic parents (8/378 vs 5/1942, *P* = 2.80×10^-4^) and the variant co-occurrence in gnomAD (8/378 vs 2/125748, *P* = 2.70×10^-19^).

None of the eight patients had pathogenic or likely pathogenic variants in the genes known to be associated with epileptic phenotypes.[30]

### Clinical features of the cases with *ZFHX3* variants

The detailed clinical characteristics of the eight unrelated cases with *ZFHX3* variants are summarized in **Table 1**. The onset age of seizures ranged from 5 months to 13 years, with a median age of onset of 5 years. The patients presented with partial epilepsy. However, two patients experienced frequent spasms (Cases 4 & 6) in the early stage of the illness, and one patient experienced nonconvulsive status epilepticus (Case 7). These three cases had neurodevelopmental abnormalities and were diagnosed as DEE. The other five cases exhibited only infrequent (yearly) partial seizures or focal-origin generalized tonic-clonic seizures (Cases 1-3, 5, and 8). Focal, multifocal, and/or diffuse epileptic discharges were recorded in their EEG (**Figure 1C-F**). The brain MRI was normal in all cases. A seizure-free status was achieved in all eight patients after prompt antiepileptic-drug treatment, including the two cases with infantile spasms, who did not use adrenocorticotropic-hormone (ACTH)/steroids (**Table 1**).

### Damaging effects of *ZFHX3* variants

The *ZFHX3* protein contains 23 zinc finger motifs and 4 homeodomains. Two variants, p.Glu807Lys and p.Ala1376del, were located in the zinc finger motifs, and other variants were scattered between zinc fingers and/or homeodomains (**Figure 2A**). The variant p.Pro3618Gln, located in the last part of the C-terminus, was repetitively identified in three of five mild cases (Cases 2, 5, & 8). Similarly, the frameshift variant (p.Pro3195LeufsTer44), which truncates the C-terminus, was also identified in a mild case (Case 3). The remaining mild case (Case 1) had two variants located in the N-terminus. The specific location of the variants in mild cases indicated a possible molecular subregional effect.[31]

**Figure 2.**
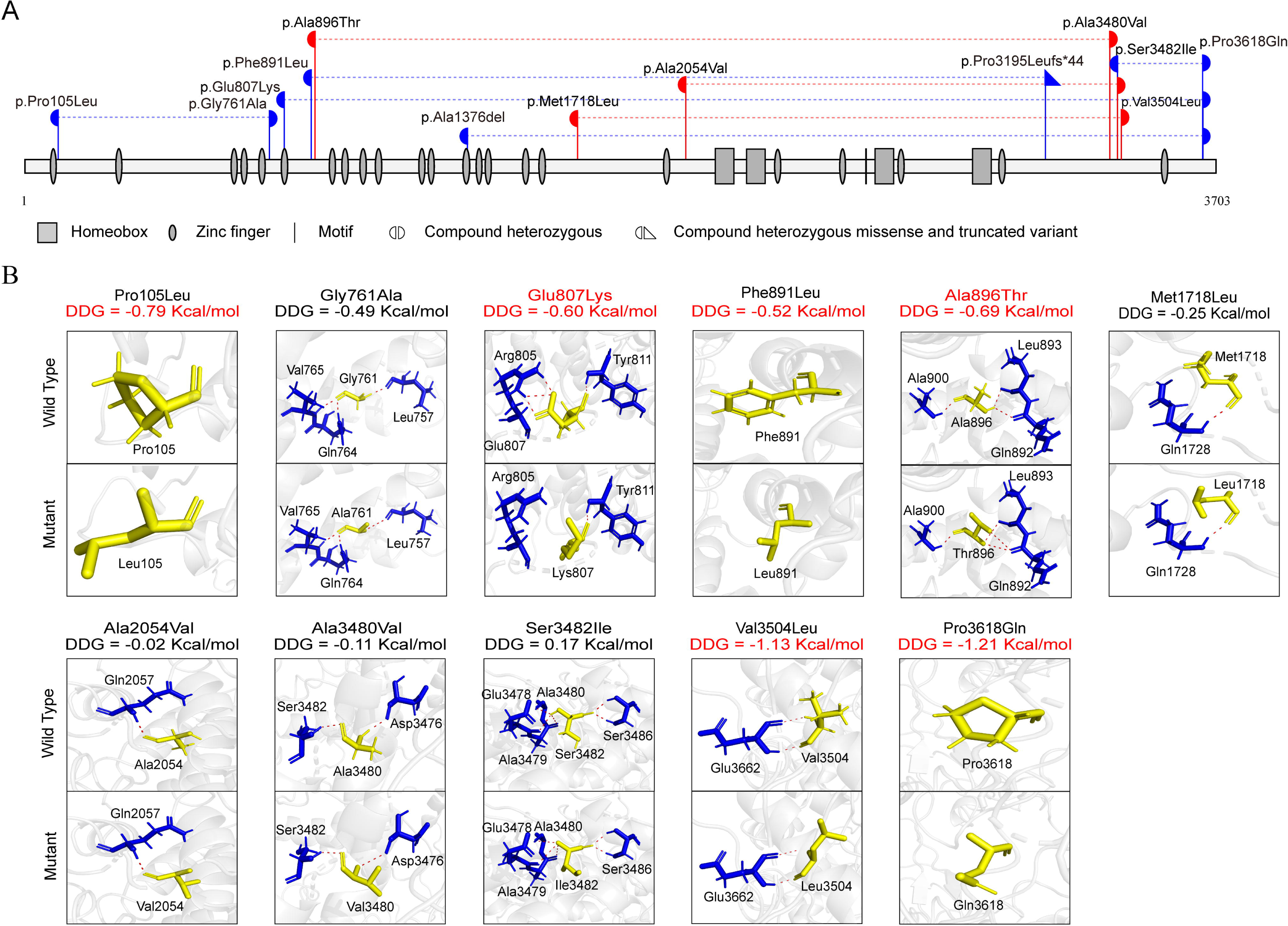
Schematic illustration of *ZFHX3* variants. (A) Schematic illustration of zinc finger homeobox protein 3 and the locations of the *ZFHX3* variants identified in this study. Two variants of the same height were a pair of biallelic variants. The red color represents cases with severe partial epilepsy with neurodevelopmental abnormalities. The blue color represents cases with mild partial epilepsy. (B) Hydrogen bond changes (red) and free energy stability changes (△△G, Kcal/mol) of the variants from the present study.

The damaging effect of missense variants was analyzed by protein modeling. Two missense variants, p.Glu807Lys and p.Ala896Thr, were predicted to alter hydrogen bonding with surrounding residues. Six variants were predicted to decrease protein stability with a LLG value less than -0.5 kcal/mol (**Figure 2B**). All the variants were predicted to be damaging by at least one *in silico* tool (**Table S3**).

### Knockdown of *Zfh2* in *Drosophila* led to increased susceptibility to seizures

To validate the association between *ZFHX3* and epilepsy, a *Drosophila* model of *Zfh2* knockdown was established, with a knockdown efficiency of approximately 66% (**Figure 3A**). The larval development of *Zfh2* knockdown flies was initially investigated. The larval body length of the *Zfh2* knockdown flies was similar to that of the *UAS-Zfh2-RNAi* control flies (**Figure 3B**).

**Figure 3.**
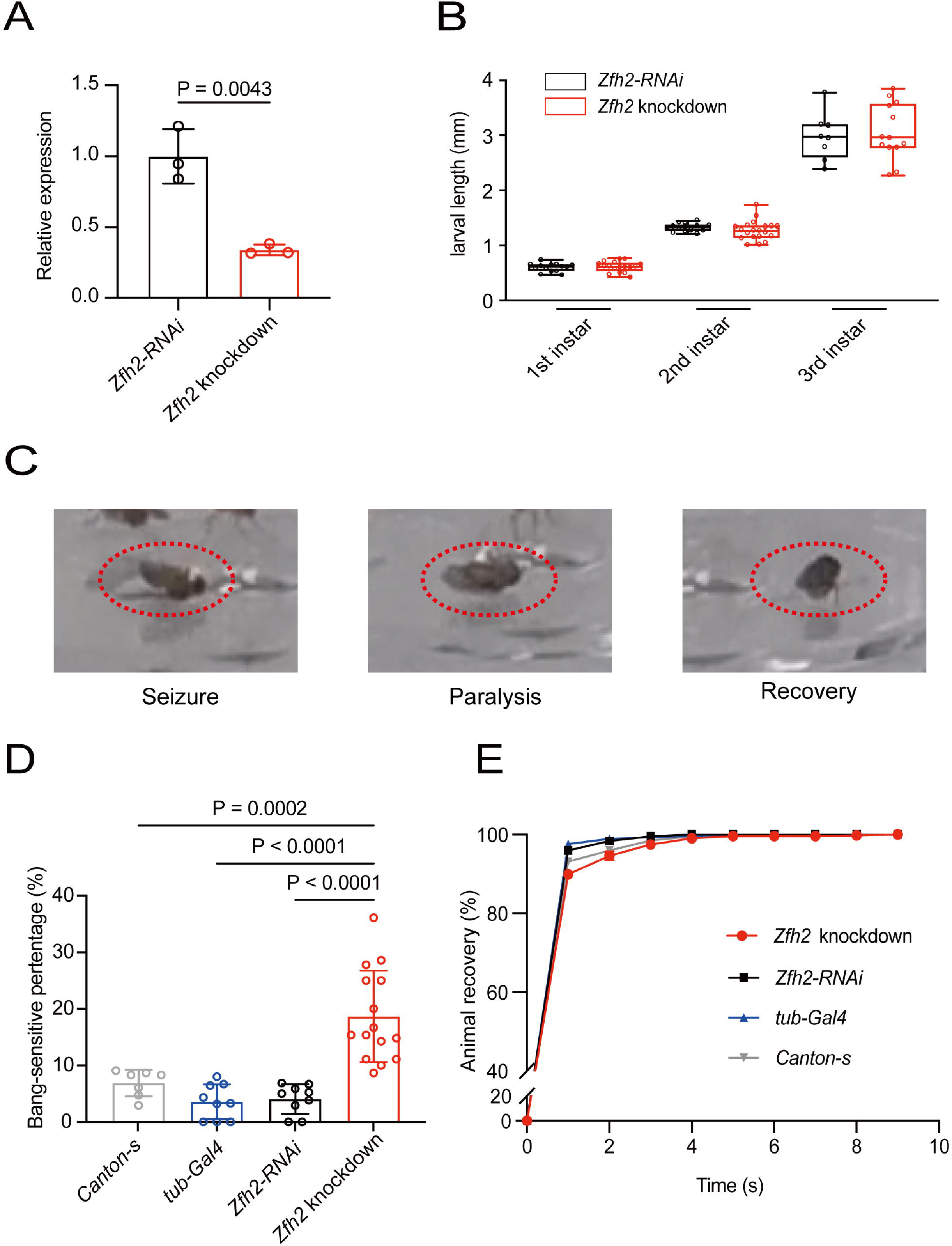
Functional studies on *Zfh2* knockdown flies. (A) Relative *Zfh2* mRNA expression levels of the knockdown flies and the *UAS-Zfh2-RNAi* controls. (B) The body length of *Zfh2* knockdown and *UAS-Zfh2-RNAi* control larvae. (C) Behavior in the Bang-sensitive test; the three phases observed in *Zfh2*-knockdown flies were seizure, paralysis, and recovery. (D) Seizure-like behaviors occurred at a higher rate in *Zfh2* knockdown flies (*tub-Gal4 > UAS-Zfh2-RNAi*) than in the *UAS-Zfh2-RNAi* line. (E) The recovery time from seizure-like behaviors of *Zfh2* knockdown flies was longer than that of the controls.

A bang sensitivity assay was performed on *Zfh2* knockdown flies and *UAS-Zfh2-RNAi* control flies to assess susceptibility to seizures. The three phases of seizure activity, seizure, paralysis, and recovery, were observed in the Bang-sensitivity test (**Figure 3C**). The percentage of seizure-like behaviors in the *Zfh2* knockdown flies was significantly higher than that of *UAS-Zfh2-RNAi* control flies [18.66 ± 2.09% (*n* = 15) vs 4.06 ± 0.87% (*n* = 9); *P* < 0.0001], *Canton-s* control flies [18.66 ± 2.09% (*n* = 15) vs 6.88 ± 0.89% (*n* = 7); *P* = 0.0013], and *tub-Gal4* control flies [18.66 ± 2.09% (*n* = 15) vs 3.56 ± 1.03% (*n* = 7); *P* < 0.0001] (**Figure 3D**). The duration of seizure-like behavior in *Zfh2* knockdown flies was also longer than that of the controls (**Figure 3E**).

The effect of *Zfh2* deficiency on the electrophysiological activity of projection neurons, which are important excitatory neurons in the central nervous system of *Drosophila*,[32] was examined (**Figure 4A, B**). The frequency of sEPSPs in *Zfh2* knockdown flies was significantly higher than that in wild-type flies [0.6312 ± 0.06 Hz (*n* = 7) vs 0.09907 ± 0.01 Hz (*n* = 7); *P* < 0.0001] (**Figure 4C)**. There was no significant difference in the amplitude of sEPSPs between the *Zfh2* knockdown flies and WT flies [1.405 ± 0.2039 mV (*n* = 7) vs 1.228 ± 0.1486 mV (*n* = 7), *P* = 0.0888] (**Figure 4D**).

**Figure 4.**
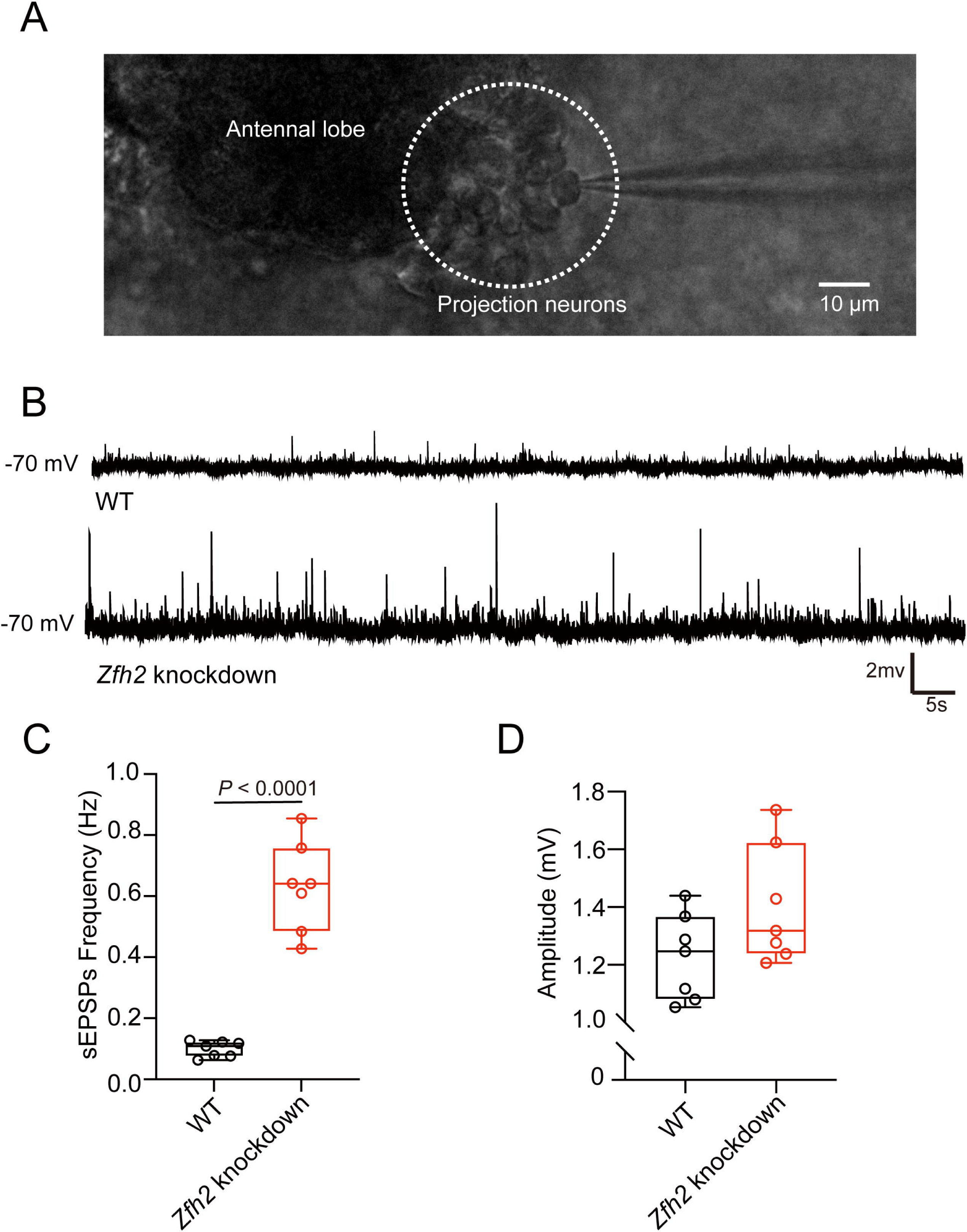
*Zfh2* knockdown induces increased neural excitability in projection neurons. (A) Whole-cell recording in the fly brain. (B) Representative traces of projection neuron sEPSPs in *Canton-s* wild-type (WT) flies and *Zfh2* knockdown flies. (C) The frequency of sEPSPs was significantly higher in *Zfh2*-knockdown flies than in wild-type flies. (D) There was no difference in the amplitude of sEPSPs between *Zfh2*-knockdown flies and wild-type flies.

### The temporal expression stage of *ZFHX3* orthologs

Infantile spasms are generally a severe form of epilepsy with a poor prognosis in most cases.[33] In this study, the two patients with early spasms presented with favorable outcomes. Recent studies have indicated that the genetic-dependent (expression) stage (GDS) is associated with the evolutional course and outcomes of illness.[14, 24] We thus analyzed the temporal expression pattern of *ZFHX3* orthologs. In *Drosophila*, the expression of *Zfh2* was high in larvae, decreased in pupae and early adults, and increased in later adults (**Figure 5A**). In mice, the expression level of *Zfhx3* was high in the fetus, decreased dramatically after birth, and slightly increased in later adults (32 weeks) (**Figure 5B**). We further analyzed the temporal expression pattern of *ZFHX3* in the human brain by using data from BrainSpan. *ZFHX3* was also highly expressed in the embryonic stage, decreased dramatically in childhood, with a nadir at approximately 10 years of age, and slightly increased at approximately 30 years of age (**Figure 5C**).

**Figure 5.**
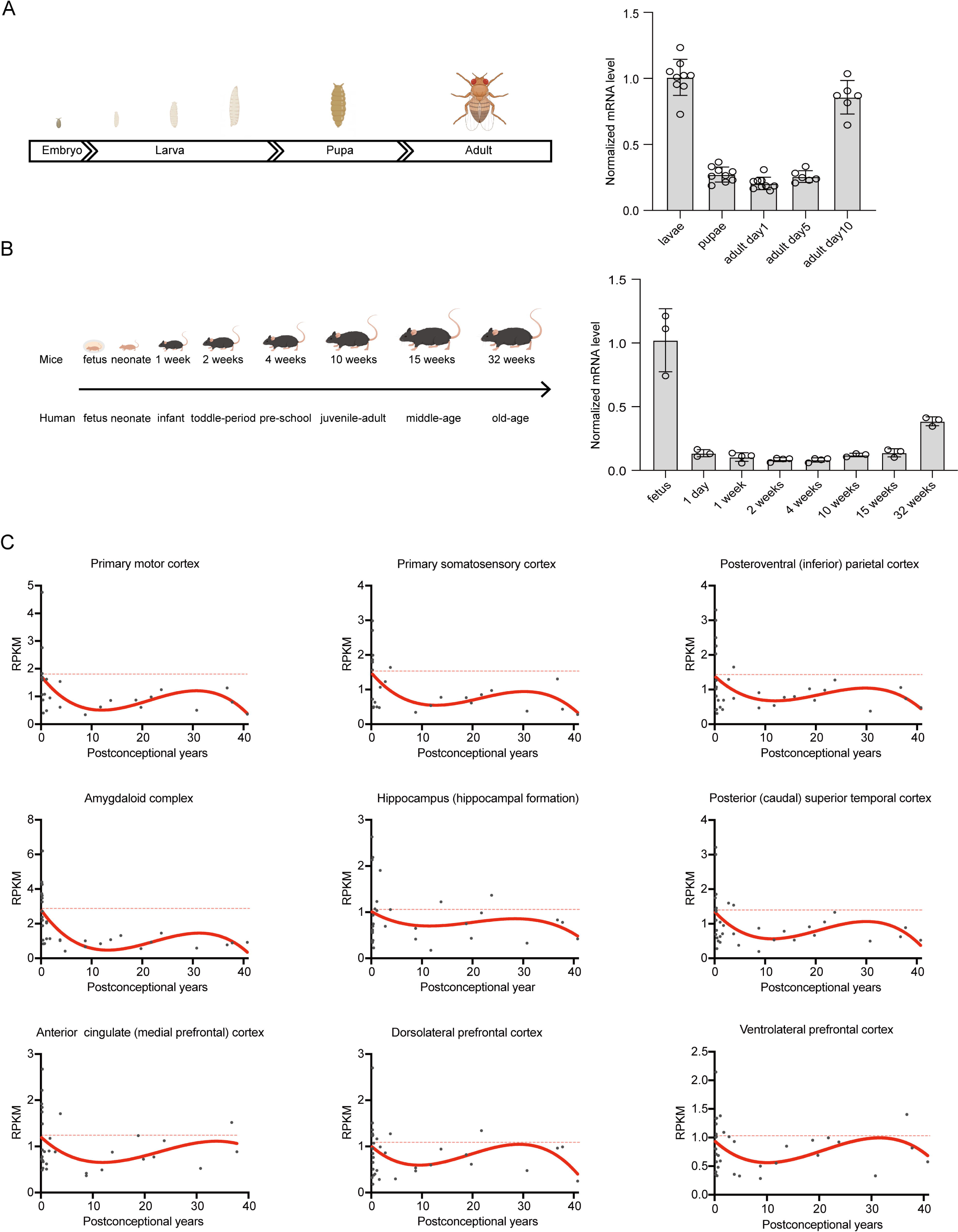
The temporal expression profile of *ZFHX3* orthologs. (A) Schematic illustration of the developmental stages of *Drosophila melanogaster* (left) and the temporal expression of *Zfh2* in flies (right). The mRNA levels of *Zfh2* were examined by RT-qPCR. The *Zfh2* mRNA levels were normalized to the mRNA levels in the third instar larvae. (B) Schematic illustration of the developmental stages of mice (left) and the temporal expression of *Zfhx3* in the frontal cortex of mice (right). The *Zfhx3* mRNA levels were normalized to the mRNA levels in fetal mice. (C) Temporal expression pattern of *ZFHX3* in the human brain. RPKM, reads per kilobase per million mapped reads.

## DISCUSSION

The zinc finger homeobox 3 gene (*ZFHX3,* OMIM *104155), also known as AT motif-binding transcription factor 1 gene (*ATBF1*), encodes a transcription factor with 4 homeodomains and 23 zinc finger motifs. In this study, we identified compound heterozygous *ZFHX3* variants in eight unrelated patients presented with partial epilepsy, including three patients diagnosed as DEE. The *ZFHX3* gene exhibited significantly higher excesses of variants in the case cohort than in the control cohort according to multiple/specific statistical analyses. Knockdown of *Zfh2* in flies increased susceptibility to seizures and abnormal firing of neurons. All patients achieved seizure-free status after prompt treatment, including the three patients with DEE. Analysis of the temporal expression profile indicated that *ZFHX3* orthologs were highly expressed in the embryonic stage and decreased dramatically in childhood, which was correlated with favorable outcomes of the patients. This study suggested that *ZFHX3* is a novel causative gene of childhood partial epilepsy and DEE.

The *ZFHX3* gene is highly conserved with homologs in flies, zebrafish, mice, and humans. It is ubiquitously expressed, especially in the developing brain. *ZFHX3* plays vital roles in multiple biological processes, including embryonic development, cell proliferation, neuronal differentiation, and neuronal death.[34-36] In mice, homozygous knockout of *Zfhx3* led to prenatal lethality with complete penetrance, while heterozygous knockout resulted in growth restriction and postnatal lethality with incomplete penetrance.[37] In zebrafish, knockdown of *Zfxh3* caused significantly increased pentylenetetrazol-induced seizures.[38] In flies, previous studies have shown that the majority of knockout/knockdown *Zfh2* lines exhibited preadult lethality (http://flybase.org/reports/FBgn0004607#phenotypes). This study identified biallelic *ZFHX3* variants in patients with partial epilepsy and DEE. The *Zfh2* knockdown in *Drosophila* led to increased susceptibility to seizures and abnormal firing of neurons. The recurrent epileptic and/or neurodevelopmental phenotypes across species supported the associations between *ZFHX3* and epilepsy and neurodevelopmental abnormalities.

*ZFHX3* is intolerant to loss-of-function (LOF) variants, as indicated by constraint indices such as the probability of being LOF intolerant (pLI),[39] the LOF observed/expected upper bound fraction (LOEUF),[39] and the probability of haploinsufficiency (pHaplo).[40] The *ZFHX3* gene has a pLI of 0.997 (> 0.9), a LOEUF of 0.147 (< 0.35), and a pHaplo of 1 (≥ 0.86), all indicating that *ZFHX3* is intolerant to heterozygous LOF variants. Considering the recurrent lethal and neurodevelopmental/epileptic phenotypes of *Zfhx3* knockout/knockdown animals, it is possible that severe *ZFHX3* deficiency, such as biallelic null variants, may cause early lethality, while moderate *ZFHX3* deficiency, such as heterozygous null variants or biallelic missense variants with moderate damaging effects, is potentially associated with epilepsy and/or neurodevelopmental abnormalities. In this study, most of the variants were located outside the functional domains and were predicted to be without hydrogen bond alterations with surrounding residues, indicating mild damaging effects, which potentially explain the pathogenesis of compound heterozygous variants. Three of the patients with a mild phenotype had the identical variant p.Pro3618Gln, which was located in the last part of the C-terminus and presented a low MAF (0.000301) (instead of absent) in the general population, indicating a potentially mild damaging effect. One patient with a mild phenotype had a frameshift variant with truncation at the C-terminus, which was inherited from the asymptomatic father and presented with a low MAF (instead of absent), thus potentially resulting in mild damage. These findings suggested a potential genotype-phenotype correlation.

The ZFHX3 protein is a transcription factor that contains 23 C_2_H_2_-type zinc fingers and 4 homeodomains and belongs to the C_2_H_2_-type zinc finger protein family, which plays an essential role in neurodevelopment.[41] Previously, *ZFHX3* de novo variants were occasionally detected in patients with neurodevelopmental disorders, including four variants in autism spectrum disorder,[42-44] three variants in developmental disorder,[45] and one variant in developmental and epileptic encephalopathy (**Table S4**),[46] suggesting a potential role in neurodevelopmental disorders. To date, 112 genes have been defined as causative genes of DEE (www.omim.org). In the present study, neurodevelopmental abnormalities were observed in three patients who were diagnosed as DEE. These data suggested that *ZFHX3* is a novel pathogenic gene of DEE.

Recently, a study suggested that *ZFHX3* binds to the promoter regions of human neural stem cells, especially those implicated in the regulation of the expression of genes in the Hippo/YAP and mTOR pathways.[47] The Hippo/YAP pathway plays vital roles in multiple stages of neuronal development;[48] while genes in the mTOR pathway, such as *TSC1*, *TSC2*, *DEPDC5*, and *SZT2*, have been identified as common genes involved in epilepsy/neurodevelopmental diseases.[6, 49, 50] Dysfunction of *ZFHX3* could disrupt the signal transduction of the Hippo/YAP and mTOR pathways and might be involved in neurodevelopmental diseases/epilepsy, which warrant further studies.

Infantile spasms is a common form of DEE that generally presents severe epilepsy with poor outcomes.[4] Clinically, approximately 33% of patients with spasms present with favorable outcomes,[33] but the underlying mechanism is unknown. Our recent study showed that GDS is associated with the evolutional course and outcomes of diseases.[14, 24] In this study, two patients with early spasms achieved seizure-free after prompt antiepileptic-drug treatment without ACTH/steroids, which were commonly used for treatment of infantile spasms but potentially with severe side effects. The genetic diagnosis thus implies a significance in precise treatment of the patients with *ZFHX3* variants. Studies on the temporal expression stage showed that *ZFHX3* orthologs were highly expressed in the embryonic stage and decreased dramatically in childhood, which is potentially one of the explanations for these favorable outcomes. Considering that the expression of *ZFHX3* slightly increased at approximately 30 years of age, long-term follow-up is needed to observe the future progression of this disease.

In conclusion, this study suggested that *ZFHX3* is a novel causative gene of childhood partial epilepsy and DEE. The disclosed genotype-phenotype correlation explained the phenotypic variation. The patients with infantile spasms achieved seizure-free after prompt treatment without ACTH/steroids, implying a significance in precise treatment of the patients. The correlation between the outcome and GDS provided an insight into the underlying mechanism of the evolutional course of the illness, potentially being helpful in the management of the patients.

## Data Availability

Raw data were generated at Institute of Neuroscience, The Second Affiliated Hospital of Guangzhou Medical University. Derived data supporting the findings of this study are available from the corresponding author on request.

## Acknowledgments

We thank the patients and their families for participating in this study. We thank Prof. Liu Ji-Yong for donating the *tub-Gal4* driver line and *Canton-s* line flies and Schrödinger (New York, NY, USA) for supporting the open-source PyMOL software.

## Contributors

W-PL, Y-HY, NH, and J-DQ designed this study; data collection was performed by M-FH, L-HL, SL, JW, Q-X Z, S-LH, D-FZ, X-RL, B-ML, H-YM, and PZ; formal analysis was performed by M-FH and SL; experiments were performed by M-FH and J-JG; grant recipients were W-PL, Y-HY, NH, and X-RL; visualization was performed by M-FH and P-YW; the first draft of the manuscript was written by M-FH, L-HL, and SL; writing-review and editing were performed by NH, Y-HY, and W-PL.

## Ethics approval statement

This study was approved by the Ethics Committee of The Second Affiliated Hospital of Guangzhou Medical University (2020-h5-49), and written informed consents were obtained from all patients or their parents. The studies adhered to the guidelines of the International Committee of Medical Journal Editors with regard to patient consent for research or participation.

## Data availability statement

Raw data were generated at the Institute of Neuroscience, The Second Affiliated Hospital of Guangzhou Medical University. Derived data supporting the findings of this study are available from the corresponding author upon request. The data are not publicly available due to privacy or ethical restrictions.

## Supplemental information

Supplemental Table S1: Primer sequences for RT-qPCR.

Supplemental Table S2: Analysis of the aggregate frequency of *ZFHX3* variants identified in this study.

Supplemental Table S3: Genetic features of the individuals with *ZFHX3* variants.

Supplemental Table S4: Previously reported *ZFHX3* variants and associated phenotypes.

Supplemental Table S5: Detailed clinical features of the individuals with *ZFHX3* variants.

