## Supplementary material for "*ZFHX3* variants cause childhood partial epilepsy and infantile spasms with favorable outcomes": Table S1

**Table S1. Primer sequences for RT-qPCR**

| | |
|---|---|
| Primers for *Drosophila melanogaster* | |
| *GAPDH*-F: | 5’-CGTCAACGATCCCTTCATCGATGTC-3’ |
| *GAPDH*-R: | 5’-CAGCACTGGCCCAGTTGATGTTG-3’ |
| *Zfh2*-F: | 5’-GCGATAACAAGAGGCAGGAGG-3’ |
| *Zfh2*-R: | 5’-CGGACACTACACGAAAAGAATGG-3’ |
| Primers for mice | |
| *GAPDH*-F: | 5’-TGTTCCTACCCCCAATGTGTC-3’ |
| *GAPDH*-R: | 5’-AAGTCGCAGGAGACAACCTG-3’ |
| *Zfhx3*-F: | 5’-GACAAACCCAGTAGCATGGAG-3’ |
| *Zfhx3*-R: | 5’-GTAGGTCTGGAGGCTGGAAAAG-3’ |
