## Supplementary material for "*ZFHX3* variants cause childhood partial epilepsy and infantile spasms with favorable outcomes": Table S2

### Table S2. Analysis of the aggregate frequency of *ZFHX3* variants identified in this study

| Identified *ZFHX3* variants | Allele count/number in this study | Allele count/number in controls of gnomAD-all populations | Allele count/number in controls of gnomAD-East Asian populations | Allele count/number in ExAC populations | Allele count/number in controls of Epi25 WES browser | Number of Homozygotes in controls of gnomAD |
|---|---|---|---|---|---|---|
| c.314C>T/p.Pro105Leu | 1/756 | 34/112352 | 25/9804 | 23/104844 | 15/66264 | 0 |
| c.2282G>C/p.Gly761Ala | 1/756 | 22/109332 | 11/9042 | 16/119178 | 10/66888 | 0 |
| c.2419G>A/p.Glu807Lys | 1/756 | 10/109406 | 0/9046 | 6/121404 | 1/66888 | 0 |
| c.2671T>C/p.Phe891Leu | 1/756 | –/– | –/– | –/– | –/– | - |
| c.2686G>A/p.Ala896Thr | 1/756 | 24/108908 | 1/9046 | 17/119382 | 2/66868 | 0 |
| c.4125_4127del/p.Ala1376del | 1/756 | 8/120262 | 8/9962 | 6/121388 | 0/66888 | 0 |
| c.5152A>C/p.Met1718Leu | 1/756 | 2/109258 | 2/9022 | 1/119740 | 1/66886 | 0 |
| c.6161C>T/p.Ala2054Val | 1/756 | 2/105822 | 1/8870 | 2/115778 | –/– | 0 |
| c.9583_9584insT/p.Pro3195LeufsTer44 | 1/756 | 8/119370 | 2/9686 | 60/118664 | –/– | 0 |
| c.10439C>T/p.Ala3480Val | 1/756 | 7/119956 | 7/9942 | 5/118166 | 0/66884 | 0 |
| c.10445G>T/p.Ser3482Ile | 2/756 | 30/119978 | 30/9938 | 25/117840 | 9/66884 | 0 |
| c.10510G>C/p.Val3504Leu | 1/756 | –/– | –/– | –/– | –/– | - |
| c.10853C>A/p.Pro3618Gln | 3/756 | 35/116270 | 35/9678 | 23/112466 | 5/66408 | 0 |
| Total | 16/756 | 182/120262 | 122/9962 | 184/121404 | 43/66888 | 0 |
| P value† | | $2.3\times10^{-13}$ | $4.36\times10^{-2}$ | $2.34\times10^{-13}$ | $3.56\times10^{-18}$ | |
| OR (95% CI) | | 14.27 (7.94-23.95) | 1.74 (0.96-2.97) | 14.25 (7.93-23.91) | 33.61 (18.85-59.94) | |

†P values and odds ratio were estimated with a 2-sided Fisher's exact test.

Abbreviations: CI, confidence interval; gnomAD, Genome Aggregation Database; ExAC, Exome Aggregation Consortium; Epi25, a whole-exome sequencing case-control study of epilepsy; OR, odds ratio
