## Supplementary material for "*ZFHX3* variants cause childhood partial epilepsy and infantile spasms with favorable outcomes": Table S3

**Table S3. Genetic features of the individuals with *ZFHX3* variants**

| Case | Coordinate | cDNA change (NM_006885) | Protein change | Inheritance | MAF | SIFT | PP2_Var | Mutation-Taster | CADD | GERP++ | phyloP | phastCons | SiPhy |
|---|---|---|---|---|---|---|---|---|---|---|---|---|---|
| **Case 1** | chr16:72993731 | c.314C>T | p.Pro105Leu | paternal | 2.74e-04 | T (0.056) | B (0.001) | D (1.000) | T (17.01) | C (5.11) | NC (0.874) | NC (0.534) | C (12.922) |
| | chr16:72991763 | c.2282G>C | p.Gly761Ala | maternal | 1.54e-04 | D (0.020) | P (0.719) | D (1.000) | T (19.17) | C (4.51) | C (3.856) | NC (0.964) | C (13.864) |
| **Case 2** | chr16:72991626 | c.2419G>A | p.Glu807Lys | paternal | 5.20e-05 | T (0.153) | P (0.994) | D (1.000) | D (24.70) | C (5.52) | C (7.858) | C (1.000) | C (19.474) |
| | chr16:72821322 | c.10853C>A | p.Pro3618Gln | maternal | 2.39e-04 | D (0.001) | P (0.669) | D (1.000) | T (17.82) | C (4.39) | C (6.706) | C (1.000) | C (16.590) |
| **Case 3** | chr16:72991374 | c.2671T>C | p.Phe891Leu | maternal | - | T (0.064) | P (0.977) | D (1.000) | D (25.30) | C (5.52) | C (9.268) | C (1.000) | C (15.677) |
| | chr16:72822591 | c.9583_9584insT | p.Pro3195LeufsTer44 | paternal | 3.93e-05 | - | - | - | - | - | - | - | - |
| **Case 4** | chr16:72991359 | c.2686G>A | p.Ala896Thr | paternal | 1.52e-04 | T (0.352) | B (0.015) | D (1.000) | T (19.79) | NC (-4.08) | NC (0.020) | NC (0.933) | NC (8.169) |
| | chr16:72821736 | c.10439C>T | p.Ala3480Val | maternal | 5.38e-05 | T (0.114) | P (0.970) | D (1.000) | D (22.80) | C (4.24) | C (7.999) | C (1.000) | C (16.983) |
| **Case 5** | chr16:72832453 | c.4125_4127del | p.Ala1376del | maternal | 3.18e-05 | - | - | - | - | - | - | - | - |
| | chr16:72821322 | c.10853C>A | p.Pro3618Gln | paternal | 2.39e-04 | D (0.001) | P (0.669) | D (1.000) | T (17.82) | C (4.39) | C (6.706) | C (1.000) | C (16.590) |
| **Case 6** | chr16:72831429 | c.5152A>C | p.Met1718Leu | paternal | 1.99e-05 | T (0.665) | B (0.009) | D (1.000) | T (13.16) | C (5.74) | C (4.166) | C (1.000) | C (16.331) |
| | chr16:72821665 | c.10510G>C | p.Val3504Leu | maternal | - | D (0.022) | P (0.537) | D (1.000) | D (21.00) | C (4.22) | C (4.744) | C (1.000) | C (16.949) |
| **Case 7** | chr16:72830420 | c.6161C>T | p.Ala2054Val | paternal | 1.85e-05 | D (0.000) | P (0.998) | D (1.000) | D (23.10) | C (5.29) | C (9.940) | C (1.000) | C (19.309) |
| | chr16:72821730 | c.10445G>T | p.Ser3482Ile | maternal | 2.47e-04 | D (0.015) | B (0.052) | D (0.950) | D (20.20) | C (4.24) | NC (1.133) | NC (0.797) | NC (8.770) |
| **Case 8** | chr16:72821730 | c.10445G>T | p.Ser3482Ile | maternal | 2.47e-04 | D (0.015) | B (0.052) | D (0.950) | D (20.20) | C (4.24) | NC (1.133) | NC (0.797) | NC (8.770) |
| | chr16:72821322 | c.10853C>A | p.Pro3618Gln | paternal | 2.39e-04 | D (0.001) | P (0.669) | D (1.000) | T (17.82) | C (4.39) | C (6.706) | C (1.000) | C (16.590) |

Abbreviation: B, benign; C, conserved; CADD, Combined Annotation Dependent Depletion; D, damaging; GERP, Genomic Evolutionary Rate Profiling; MAF, minor allele frequency from general population of gnomAD; NC, nonconserved; P, probably_damaging; PP2, Polyphen2_HVAR; phastCons, Phylogenetic Analysis with Space/Time models conservation scoring and identification of conserved elements; phyloP, Computation of p-values for conservation or acceleration, either lineage-specific or across all branches; SIFT, Sorting Intolerant From Tolerant; T, tolerable.
