## Supplementary material for "*ZFHX3* variants cause childhood partial epilepsy and infantile spasms with favorable outcomes": Table S4

**Table S4. Previously reported *ZFHX3* variants and associated phenotypes**

| Case | Variants (NM_006885) | Original | MAF | Phenotypes | References |
|---|---|---|---|---|---|
| Case 1 | c.2532C>A/His844Gln | *De novo* | - | ASD | Wang (2016) |
| Case 2 | c.3808C>T/His1270Tyr | *De novo* | - | ASD | Hashimoto (2016) |
| Case 3 | c.4857dupA/His1620Thrfs*42 | *De novo* | - | ASD | Wang (2016) |
| Case 4 | c.9622dupC/Gln3208Profs*31 | *De novo* | - | ASD | Kosmicki (2017) |
| Case 5 | c.6880C>T/Arg2294* | *De novo* | - | DD | McRae (2017) |
| Case 6 | c.8993G>A/Arg2998Gln | *De novo* | - | DD | McRae (2017) |
| Case 7 | c.3035_3036delAC/Tyr1012Serfs*34 | *De novo* | - | DD | Turner (2019) |
| Case 8 | c.7562C>T/Ala2521Val | *De novo* | - | DEE | Epi4K (2013) |
